# The Impact of the Harmonised Education Tariff on Faculty Development and Teaching Capacity in Undergraduate GP Education: A Qualitative Interview Study

**DOI:** 10.1101/2025.11.24.25340864

**Authors:** Catharina Savelkoul, Sophie Park

**Author notes:** Corresponding author, Miss Catharina Savelkoul, PhD Candidate, University of Oxford, Nuffield Department of Primary Care Health Sciences, Radcliffe Primary Care Building, Oxford, OX2 6GG, UK,. Catharina Savelkoul is a DPhil student in Primary Health Care at the University of Oxford, researching career choice into general practice. Sophie Park is a GP and Professor of Primary Care and Clinical Education at the University of Oxford.

## Abstract

**Background:** Until 2022, undergraduate general practice (GP) placements in England received substantially less funding than hospital placements. The harmonised undergraduate medical tariff established equal funding for primary and secondary care education. There is no published evidence on its impact.

**Aim:** To examine how the harmonised tariff impacted the opportunities and challenges in recruiting and retaining teaching faculty, and in coordinating, delivering, and sustaining undergraduate GP teaching in England.

**Design and setting:** A qualitative interview study with Heads of Undergraduate GP Teaching (HUGPTs) at medical schools across the UK.

**Method:** We conducted twelve semi-structured online interviews with HUGPTs from 12 different UK medical schools. We used reflexive thematic analysis: we coded inductively and iteratively, refining the topic guide as new themes emerged from early interviews.

**Results:** Six themes were identified regarding the impact of the harmonised tariff: budget control and organisational change; curriculum changes; placement capacity; teaching quality and assurance; impact on student experience; and the use of accountability frameworks, including a formal use-of-funds policy (‘Annex C’) and a reporting tool.

**Conclusion:** This study demonstrates that the harmonised tariff led to substantial improvements in undergraduate GP teaching and recruitment. Locating budgetary decision-making authority with HUGPTs increased transparency and budget allocation, and enabled the expansion of GP teaching in the curriculum and recruitment to the central GP team. Better remuneration improved recruitment and retention of placement providers and was perceived to increase the quality of teaching.

## Introduction

Funding for undergraduate medical placements in general practice has historically lagged behind that for hospital placements. The Service Increment for Teaching (SIFT) system, introduced in 1976 to fund clinical teaching in NHS hospitals, initially excluded general practices [1,2]. SIFT was only extended to GP teaching in 1995 at a token rate of £12.50 per student per half-day [1,2]. Under this legacy model, medical schools delivered around 15% of clinical curriculum hours in general practice but received only about 7% of clinical placement funding for primary care [3].

By the 2010s, this arrangement was widely regarded as outdated and inequitable [1,4–6]. The number of academic GP departments had halved since 2002 and the duration of GP placements per student was stagnating or falling [3]. This was concerning given the Department of Health’s target for 50% of graduates to enter general practice [5,6]. Adequate undergraduate GP experience is known to influence career choice, so insufficient and underfunded placements threatened future GP recruitment [7–9]. There was an urgent need for a new funding model for undergraduate primary care education in England.

In 2019, a national costing exercise by the Department of Health’s Primary Care Education Working Group found GP teaching costs were comparable to secondary care, yet GP practices were reimbursed at barely half this level [1]. This led to extensive negotiations through the National Tariff Advisory Group, bringing together the Department of Health and Social Care in England (DHSC), Health Education England and other key stakeholders [6].

In March 2022, the DHSC introduced guidance for a new undergraduate tariff providing consistent national funding for clinical teaching across all settings, with GP teaching funded on a par with secondary care for the first time [6,10]. Detailed in Annex C of the 2022-23 Education and Training tariff and informed by the National Tariff Advisory Group [6,11], the policy introduced two organisational changes: (1) the HUGPT became the budget holder for all undergraduate primary care funding devolved by NHS England (NHSE); and (2) HEIs must submit an annual accountability report (piloted in 2022/2023) [12] detailing the HUGPT, the total funding received, and a full reconciliation of its use.

The new undergraduate tariff was expected to improve undergraduate GP teaching by introducing a *‘bespoke funding model*’ [6,p.257] that recognises the unique organisational requirements of GP teaching. It also sought to formalise the role of HUGPTs by granting ‘*a level of agency commensurate with their responsibilities*’ [6,p.258] and tools to oversee spending decisions. However, there is no published empirical evidence on its impact. This study addresses this gap by examining how the harmonised tariff has impacted the opportunities and challenges of coordinating, delivering and sustaining undergraduate GP placements in England.

This paper is both timely and necessary; in March 2025, the UK government announced that NHSE will be abolished, with its functions, including oversight of undergraduate primary care education, transferring to the DHSC within two years [13]. High-quality undergraduate GP teaching is also critical to delivering the 10-Year Health Plan’s mandate to shift care to the community and achieve the target of 50% of graduates entering general practice [5–6,14]. This paper therefore examines the successes and remaining challenges of the current funding model to inform its transition and future sustainability.

## Method

### Study design and participants

We conducted qualitative interviews with HUGPTs, typically senior academic GPs, educationalists, and members of the SAPC National Heads of Teaching Committee, who lead the central GP teaching teams (CGPTs) at UK medical schools. Terminology for this role and the academic ‘home’ of GP teaching teams vary: some are based in central medical school departments, but most sit within departments of primary care. Invitations were distributed through the SAPC network; HUGPTs, each from a different UK medical school, expressed interest and were recruited.

The sample was diverse across both institutions and individuals, representing ∼30% of the total eligible group [15]. Medical schools varied in the amount and structure of GP teaching (e.g. time and timing of placements), geographical setting, and whether they were older or newer institutions (see Table 1). HUGPTs differed in time in role (<6 months to >10 years), gender (5 female, 7 male), and specific responsibilities for coordinating GP teaching.

**Table 1.** Characteristics of participating medical schools: overview of the 12 UK medical schools included in the study.

|  | Medical school | Russell group | Medical School est.* | Cohort size** | Course | Teaching style (overall) |
| --- | --- | --- | --- | --- | --- | --- |
| English Medical Schools | i | X | New | Medium | Problem-based | PBL-based |
|  | ii | X | New | Medium | Integrated | Mixed |
|  | iii | ✓ | New | Medium | Problem-based | PBL-based |
|  | iv | ✓ | Established | Medium | Integrated | Mixed |
|  | v | ✓ | Established | Large | Integrated | Case-based |
|  | vi | ✓ | Established | Large | Integrated | Mixed methods |
|  | vii | ✓ | Established | Medium | Traditional | Traditional |
|  | viii | ✓ | Long-established | Large | Integrated | Mixed |
|  | ix | ✓ | Long-established | Large | Problem-based | PBL-based |
|  | x | ✓ | Long-established | Large | Integrated | Mixed |
| Non-English UK medical school | xi | ✓ | Long-established | Medium | Problem-based | PBL/CBL |
|  | xii | ✓ | Long-established | Medium | Integrated | Mixed |

\* Medical School Est.: New = founded post-2000, Established = founded 1950-1999, Long-established = founded pre-1950
\*\* Cohort size: Medium = <300 students per year, Large = >300 students per year

### Data collection

The University of Oxford Medical Sciences Interdivisional Research Ethics Committee (MS IDREC) reviewed the project and determined that it constituted a service evaluation not requiring formal ethical approval; the study was conducted in accordance with the Declaration of Helsinki and institutional research governance standards, with no patient or identifiable clinical data collected. Semi-structured interviews were conducted between June-August 2025 via Microsoft Teams, each lasting 30-60 minutes. An initial topic guide (Box 1) provided structure and consistency while allowing for new themes to emerge as participants discussed changes resulting from the harmonised tariff. The topic guide was designed to reflect our research questions and iteratively refined as interviews and preliminary analysis progressed.

Interviews continued on a rolling basis until 12 had been completed within the project timeframe, by which point major themes were repeated and thematic saturation was judged sufficient. Verbal informed consent to participate, including permission to record, was obtained at the start of each interview and documented. Interviews were audio-recorded, transcribed, anonymised, and cross-checked for accuracy against the original recordings. Data collected was stored securely on encrypted University servers in compliance with the institutional data governance requirements.

### Data analysis

The interview transcripts formed the data for analysis. Data analysis followed an inductive and iterative approach; we analysed the data using Braun and Clarke’s reflexive thematic analysis (RTA) [16], applying the ‘practical’ variant suited to applied health and education research [17]. The initial set of codes was guided by the topic guide. Transcripts were read and re-read, and we inductively generated new codes from the topics raised by the HUGPTs during interviews. We then developed the key themes and finalised the thematic coding tree (Table 2) through team discussions. The study and initial topic guide were pre-registered on the Open Science Framework, and all coding and analysis were conducted in NVivo 15. This study is reported in accordance with the Standards for Reporting Qualitative Research (SRQR), and the completed checklist is provided as Supplementary File 1.

**Table 2.** Theoretical lenses applied to post-harmonised tariff changes in undergraduate GP education: mapping findings to PAT, RDT and Institutional theory.

| Theory | What it explains | Evidence |
| --- | --- | --- |
| **Principal-Agent Theory (PAT)**<br>(Jensen & Meckling, 1976; Ross, 1973) | How reducing **information asymmetry** and granting HUGPTs budget sign-off aligned spend with intended primary care educational uses; strengthened monitoring/QA | *Theme 1, subtheme 1:* mechanics of control and oversight; HUGPTs describing new budget oversight |
| **Resource Dependence Theory**<br>(RDT) (Pfeffer & Salancik, 1978) | How **access to and control** of tariff funds **shifted dependencies** and leverage | *Theme 1, subtheme 2: Themes 2-3 -* leverage; HUGPTs reporting ‘more influence, power and control’ with central finance |
| **Institutional Theory** (DiMaggio & Powell, 1983; Scott, 1995/2001) | How tariff parity acted as a **legitimacy signal**, improving GP education’s status (challenging the hidden curriculum) | *Theme 1, subtheme 3:* cultural/legitimacy shift; HUGPTs noting ‘the narrative matches the pay.’ |

**Table 3.** Recommendations to strengthen and future-proof the undergraduate primary care funding model.

|  | *Recommendation* | *Action* |
| --- | --- | --- |
| 1 | Ensure information symmetry, transparent oversight and accountability | Continued use of **Annex C** and an external **reporting tool** will be crucial to maintaining consistency and transparency in operationalisation across medical schools. |
| 2 | Maintain formal budget-holder status | Ensure **ongoing access** to and **control** of primary care tariff **budgets** by those responsible for this teaching, as the designated budget holders, to support advocacy, agency and leverage within medical schools. |
| 3 | Align budget allocation with responsibilities and QA | Support **interrelationships** between **budget allocation** and **responsibilities** to address elements of the hidden curriculum and support future equity across undergraduate clinical education. |

## Results

A total of 12 interviews were conducted with HUGPTs from medical schools across England, including two comparative cases from non-English UK institutions. Data saturation was reached when no new themes emerged from additional interviews. Six key themes were identified regarding the impact of the harmonised tariff: budget control and organisational change; curriculum change; placement capacity; teaching quality and assurance; impact on student experience; and the use of accountability frameworks.

### Budget control, transparency and organisational change

Before the tariff, many HUGPTs reported limited oversight and control over how much funding was devolved to primary care or how it was spent within their institution. Under the harmonised tariff, Annex C guidance formally designates the HUGPT as the budget holder for undergraduate primary care funding.

#### Greater transparency and oversight

Nearly all participants reported that this change improved transparency in how undergraduate primary care funding is and should be allocated.

> ‘It is now much more transparent with the medical school about the money coming in being spent appropriately on primary care education.’ - HUGPT 2

#### Gaining formal control of the budget

Participants noted that being formally designated as the budget holder strengthened their ability to advocate for the needs of undergraduate primary care education within their medical school:

> ‘One of the outcomes [of the harmonised tariff] is more power and control to the HUGPT in the local financial arrangements … they can argue the case when they need more staff and more resources in a way that didn’t used to be possible.’ - HUGPT 12

Participants also reported that this new arrangement ensured the appropriate allocation and use of funds for GP teaching, as one explained:

> ‘Without the HUGPT’s sign off that the money’s been spent appropriately … it shouldn’t be released to other people.’-HUGPT 1

#### Institutional cultural change

There is substantial evidence of a persistent hidden curriculum [4,19–20] in UK medical schools, which portrays hospital specialties as more prestigious while denigrating general practice. Within this context, some participants observed that the funding parity challenged this prevailing cultural divide:

> ‘There’s just such value to having a harmonised tariff … it goes back to that “Are you going to be just a GP or are you going to specialise?” … the harmonised tariff goes some way in making us feel that we are equivalent to secondary care.’ - HUGPT 12

The harmonisation of the tariff between primary and secondary care acted as a *legitimacy signal* [18], indicating that undergraduate education in general practice is just as important as hospital-based teaching:

> ‘To have a funding model where you have apparent parity between primary care and secondary care … is very clearly a very progressive step.’ - HUGPT 4

### Curriculum expansion and educational innovation

Before the harmonised tariff, undergraduate GP teaching had plateaued or, in some cases, declined, with the amount of teaching per student falling in several medical schools [3,22]. Addressing this stagnation became a priority; the *By Choice - Not by Chance* [4] report called for better and more integration of GP education into curricula.

Several HUGPTs described how the formal designation as budget holder enabled them to achieve this curriculum expansion because the shift in resource control [22] reduced their dependence on the central medical school and allowed them to align funding more closely with the priorities for GP education.

#### Expansion of GP teaching within the curriculum

Several participants indicated that the harmonised tariff served as a catalyst for expanding GP teaching, addressing the ‘*lamentable structural inequality*’ [23,p.164] where the GP footprint in curricula was not matched by funding. For the first time, allocations were proportionate to the curriculum footprint, providing the leverage to plan, resource, and deliver expanded undergraduate GP teaching:

> ‘[The harmonised tariff] was really helpful because we were doubling up the amount of time students had in GP placement.’ - HUGPT 5

Not all participants had yet used the new tariff for curriculum change, although some described plans to do so:

> ‘The amount of clinical [GP] exposure is falling behind other institutions … there has been a lot of discussion about how we could use tariff money to facilitate that increase.’ - HUGPT 8

#### Central GP team (CGPT) growth

The scale, heterogeneity, and dispersed nature of UK primary care mean that, in most schools, key elements of undergraduate GP placements are delivered centrally by a medical school GP teaching team (CGPT). These include major placement management and coordination (often across 100+ practices), alongside financial, quality, and planning processes. Many schools also provide centrally delivered clinical teaching by GPs [4]. The tariff supports these activities [12]. Some participants reported that the additional funding provided through the new funding model facilitated recruitment to the CGPTs, with previously small and under-resourced teams reliant on ad hoc staff now able to grow:

> ‘We’ve historically been a very small team, underfunded in terms of administration support and clinical teachers, and so we were able to expand the team.’ - HUGPT 1

#### Development of educational initiatives

New models of delivery and innovative approaches are needed to prepare students for the changing healthcare system [4,14]. The harmonised tariff provided the means to develop and implement new primary care educational initiatives:

> ‘We couldn’t be as creative as we can be now … we’ve got new initiatives such as the clinical humanities project or the simulated clinics.’ - HUGPT 6

### Placement capacity

Consistent with the aim of establishing a ‘*bespoke funding model*’ [6,p.258] for undergraduate GP education, the harmonised tariff enabled several medical schools to increase their payments to teaching practices, bringing reimbursement closer to the true costs of delivering placements [1]:

> ‘[The harmonised tariff] meant that I could pay my GP teachers that are delivering placement activity better.’ - HUGPT 3

#### Recruitment of new providers

Participants consistently reported that the harmonised tariff improved their ability to recruit new teaching practices, whereas previously they had struggled:

> ‘I have noticed that [practices] are now saying yes.’ - HUGPT 3

> ‘We were able to add in funding to encourage practices who were wavering to and also recruit a couple of new quite big practices.’ - HUGPT 1

This aligns with existing evidence that inadequate remuneration has long been a barrier to recruitment of teaching practices [21,24].

#### Retention of existing providers

Several participants noted that, before the new funding model, many practices considered withdrawing as previous financial arrangements were no longer viable; the new tariff helped retain them:

> ‘We did get [practices] saying “we can’t afford to do this anymore”. That hasn’t happened once since the new tariff came in.’ - HUGPT 2

#### Ongoing concerns over capacity

Participants raised concerns about future placement capacity, citing rising medical student intakes, new medical schools [25–28], and increased training demands from postgraduate GP trainees and allied health professionals; together these pressures risk creating competition:

> ‘Many new medical schools are going to be competing for the same placements and allied healthcare professionals … wanting to have placement activity in general practice.’ - HUGPT 3

At one medical school, the harmonised tariff was reported to have mitigated some of these concerns:

> ‘We were quite worried about another medical school to be in our area … that was why we’ve tried to increase our local practices … we’re almost a bit too successful … if we did have an expansion of medical students … we’d probably be OK.’ - HUGPT 12

### Improving authentic teaching quality and assurance

#### Tutor engagement

Participants perceived that change in remuneration improved tutor satisfaction and, in some cases, the quality of teaching:

> ‘[The tariff] made the GP tutors happier because … they’re getting what they deserve.’ - HUGPT 6

> ‘[GP tutors] do a better job if they feel they’re being rewarded appropriately for it.’ - HUGPT 2

#### Quality assurance

As designated budget holders, HUGPTs now have oversight of tariff allocation and can directly link funding decisions to quality assurance processes. This integration allows them to leverage change in a timely and effective way:

> ‘[The tariff] gives us more clout … to ensure quality… if a practice is giving lots of self-directed learning sessions, we can say, “Hang on. You’re getting paid this much… you need to be giving them more teaching”.’ - HUGPT 12

### Impact on student experience

Teaching activity is always balanced, integrated and delivered alongside clinical care. The funding of teaching, therefore, has to ensure that both can flourish in synergistic ways. Students were reported to feel more valued by practices, as they were perceived to be generating appropriate income for the practice rather than being a burden.

#### Perceptions of general practice

Several participants suggested that improved remuneration and tutor engagement led to more positive student experiences and attitudes towards general practice:

> ‘[The harmonised tariff] has improved positivity towards general practice from our students.’ - HUGPT 5

However, some HUGPTs were careful not to overclaim on this point, until further evidence is available:

> ‘I think it would be quite tenuous to make a direct link … because there are so many factors.’ - HUGPT 4

#### Practical benefits for students

With more practices willing to teach following the tariff, several schools have been able to improve placement logistics for students:

> ‘I have been able to move [the placements] much more closely because local practices have now said we’ll take students.’ - HUGPT 3

Some HUGPTs used the harmonised tariff to act on student feedback:

> ‘We’re providing the tariff funding now for a full day [in the GP surgery] because students were complaining that … half a day is not enough.’ - HUGPT 9

### Accountability frameworks

#### Annex C as guidance

Guidance was set out in Annex C [11] of the 2022-2023 education and training tariff document. Participants consistently found this guidance document very helpful for clarifying appropriate tariff use for themselves and in negotiations with colleagues to define the boundaries of primary care funding. Many described using Annex C as a reference tool in discussions about the use of the primary care tariff, as it provides both *internal clarity* and *external legitimacy*.

> ‘[Annex C] has been useful because it means I can say yes and no to things.’ - HUGPT 3

#### Use of the reporting tool

External oversight by a national body (currently NHSE, moving to DHSC) through the reporting tool was seen as necessary to keep the operationalisation of the tariff fair and consistent across medical schools. Piloted by eight HUGPTs in 2022/2023, the tool was later rolled out nationally. Most have prepared it in anticipation of an NHSE request, although no formal submission has yet been sought.

Nevertheless, many participants emphasised the tool’s importance and expressed that reporting should stay mandatory in future to ensure that budget allocation was compliant with the requirements of Annex C.

> ‘[The reporting tool] makes sure that medical schools are carrying out the right processes and that heads of GP teaching are involved.’ - HUGPT 6

Consistent with Annex C, several HUGPTs emphasised the importance of regular tariff meetings with medical schools to review expenditure and ensure alignment with national guidance.

## Discussion

### Summary

High-quality undergraduate GP teaching is critical to meeting the plans outlined in the 10-Year Health Plan [14] to shift care from hospitals to the community and to achieving the target for 50% of graduates to enter general practice [5,6]. With NHSE set to be abolished and its functions transferred to the DHSC within two years [13], clear evidence of what has worked is crucial to support the appropriate continuation and strengthening of these funding arrangements in the future. This paper shows how this important change in funding model has facilitated substantial and measurable improvements in the organisation, delivery and quality assurance of undergraduate primary care medical education across England, through the following mechanisms:

Reallocating budgetary decision-making authority with HUGPTs increased transparency and repositioned the financial resources and decision-making power [22] with those directly responsible for delivering teaching. HUGPTs reported that this change reduced their dependence on the central medical school and enabled them to align funding more closely with the priorities for primary care education. It also reduced information asymmetry; while some HUGPTs already had oversight, the formal designation as budget holder ensured that all now have full visibility and accountability, directing allocated funds specifically to primary care education and reducing the risk of these funds being absorbed into other medical school budgets [29].

In turn, this reallocation [22] and greater transparency [29], enabled many HUGPTs to expand and better integrate GP teaching in the curriculum, grow the central GP team (CGPT) and introduce new educational initiatives. In several schools, this also allowed for increased payments to teaching practices. Participants reported that better remuneration improved recruitment and retention of placement providers which was perceived to increase the quantity and quality of clinical teaching. As Barber et al. [24] showed, practices that feel recognised and supported integrate teaching into the fabric of care; where support is thin, teaching drifts to the margins. The harmonised tariff matters, then, not only as a funding reform but as a relational one. Further research is needed to understand whether and how changes in remuneration influence teaching quality and student career intention. Together, these changes led to more and better delivery of GP teaching (*see* Figure 2).

**Figure 1.**
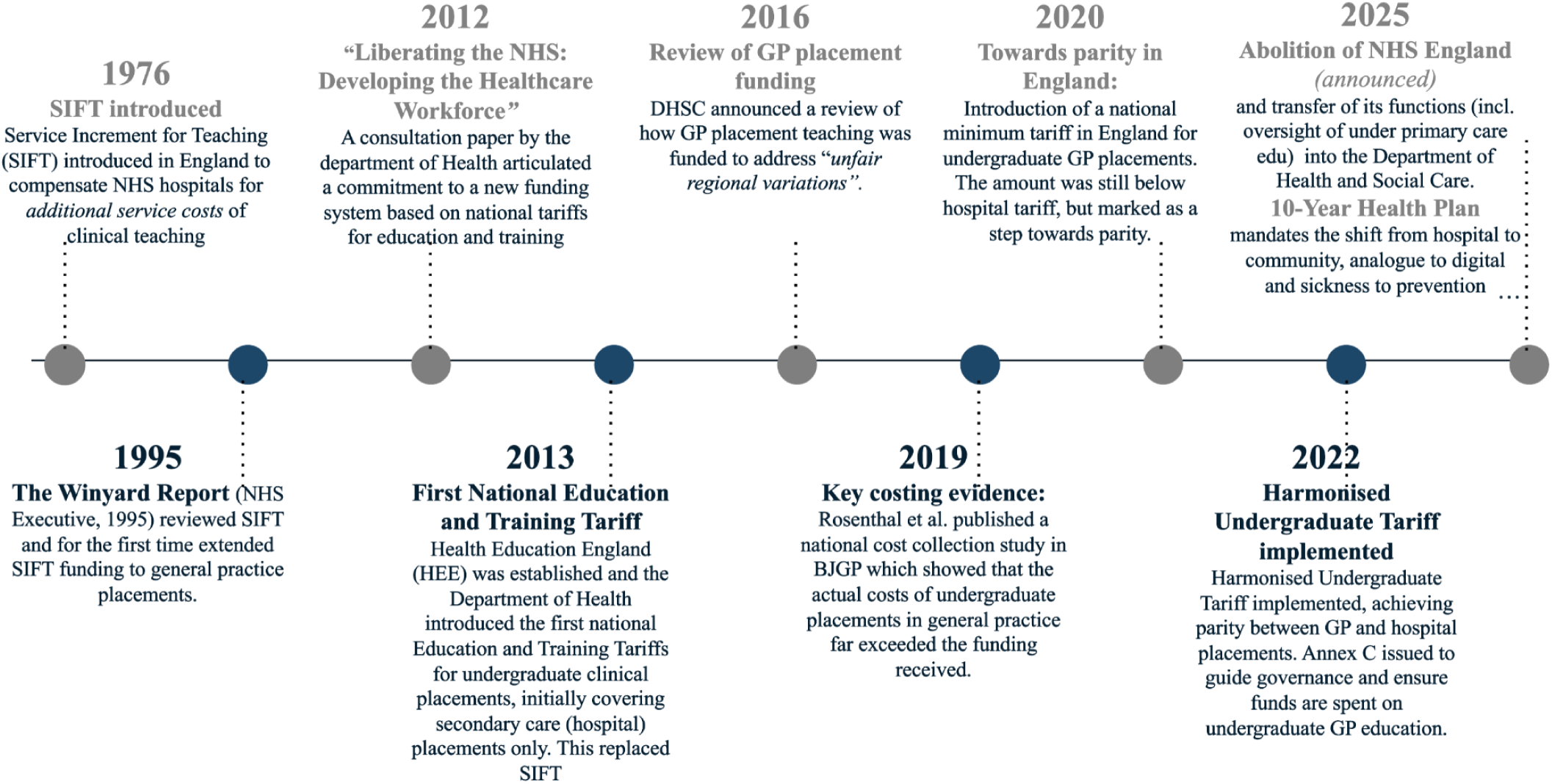
Key changes in funding and tariff arrangements for undergraduate general practice placements leading up to the harmonised undergraduate tariff, 1976-2025 (figure created by authors)

**Figure 2.**
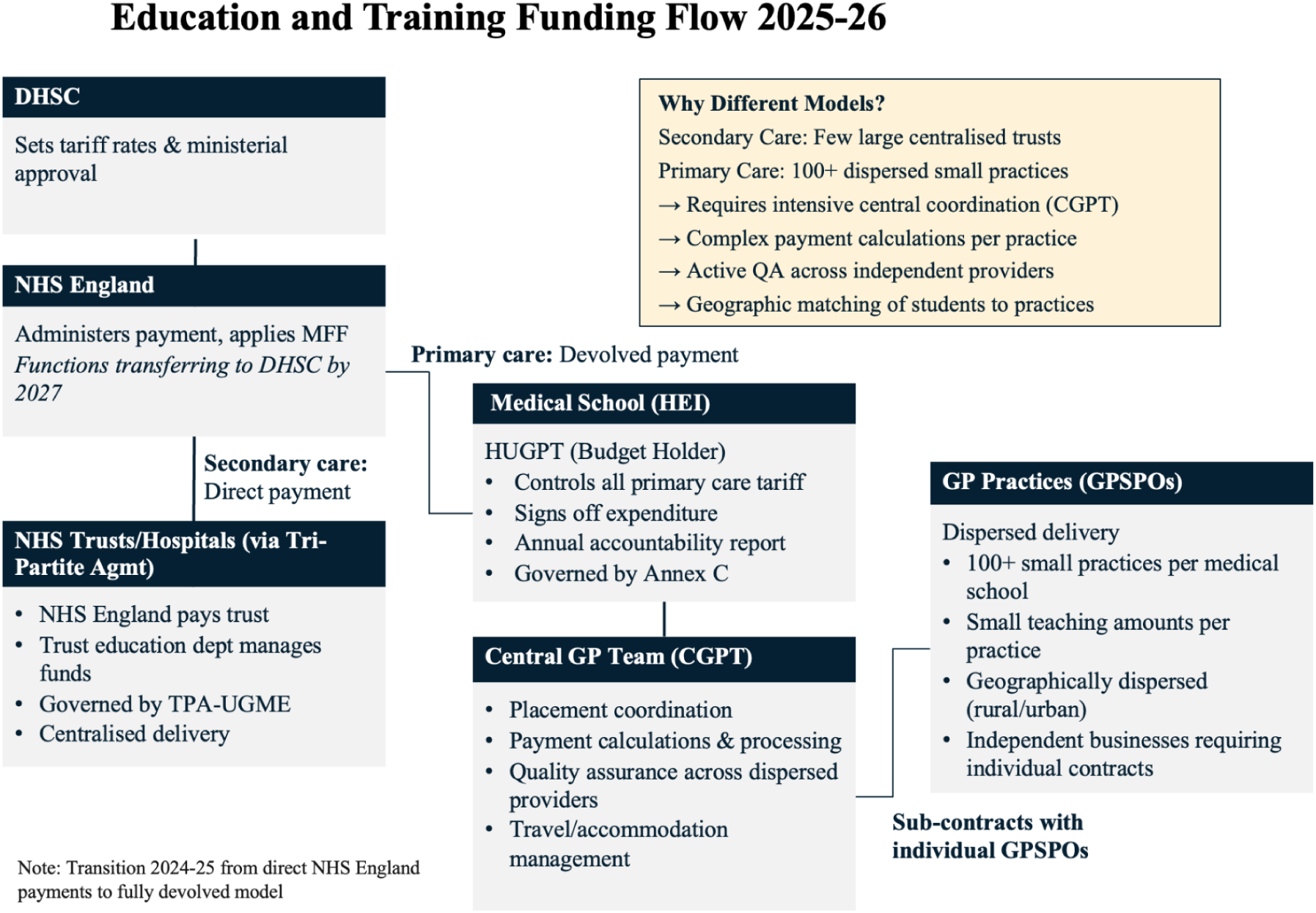
Education and Training Funding Flow (2025-2026)

**Figure 3.**
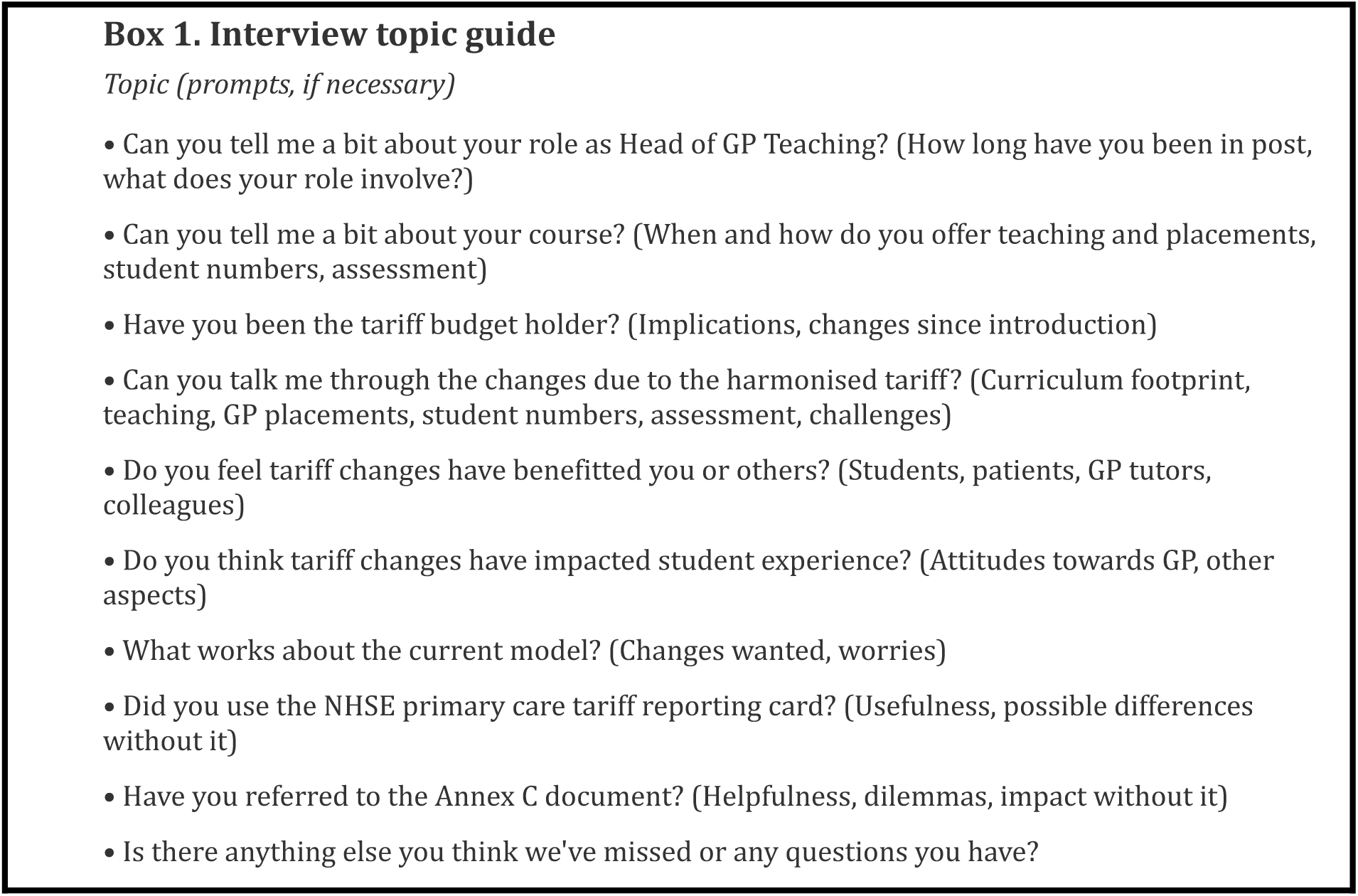
Interview topic guide

**Figure 4.**
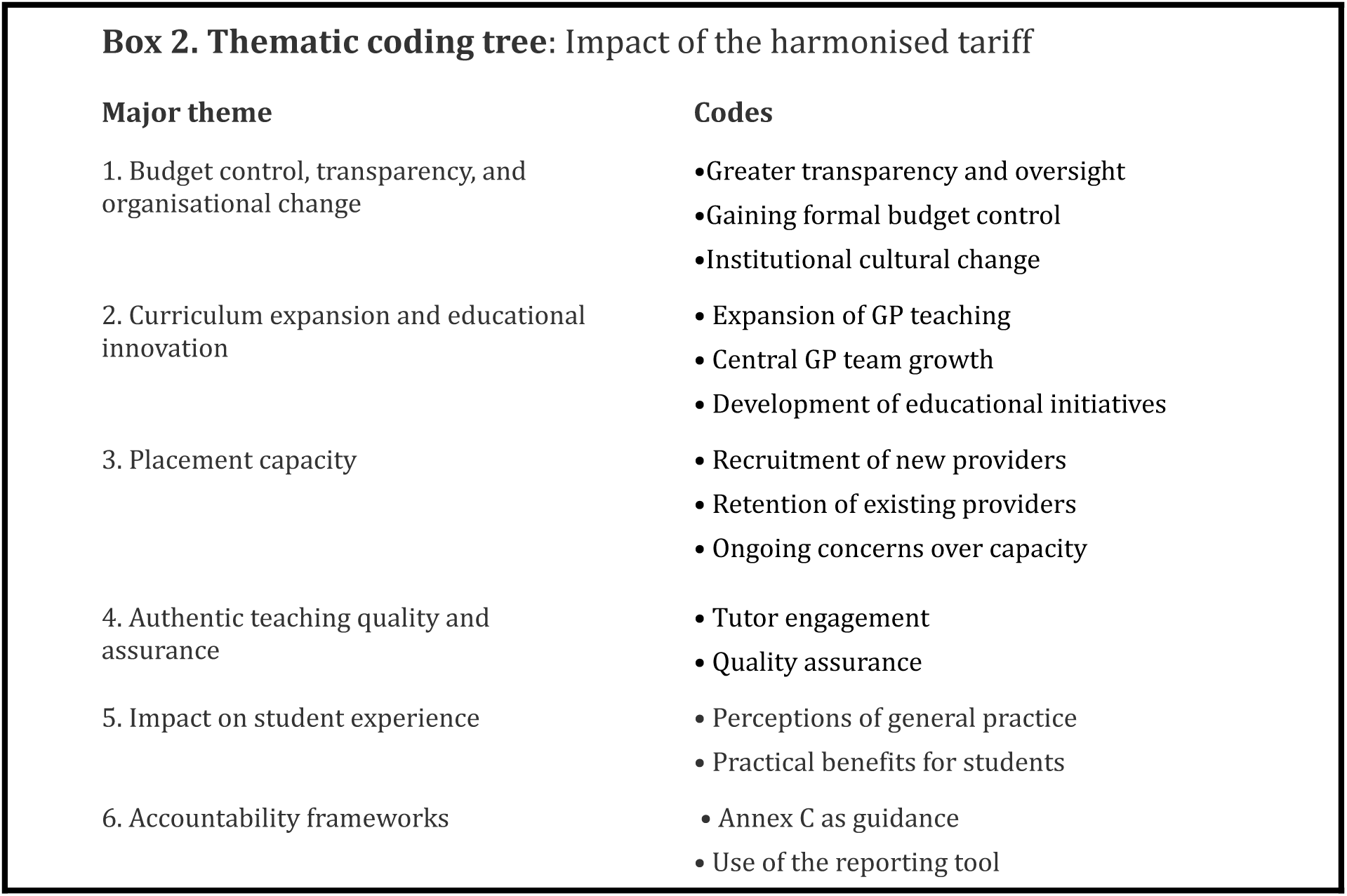
Thematic coding tree illustrating major themes and sub-themes identified in interviews.

**Figure 5.**
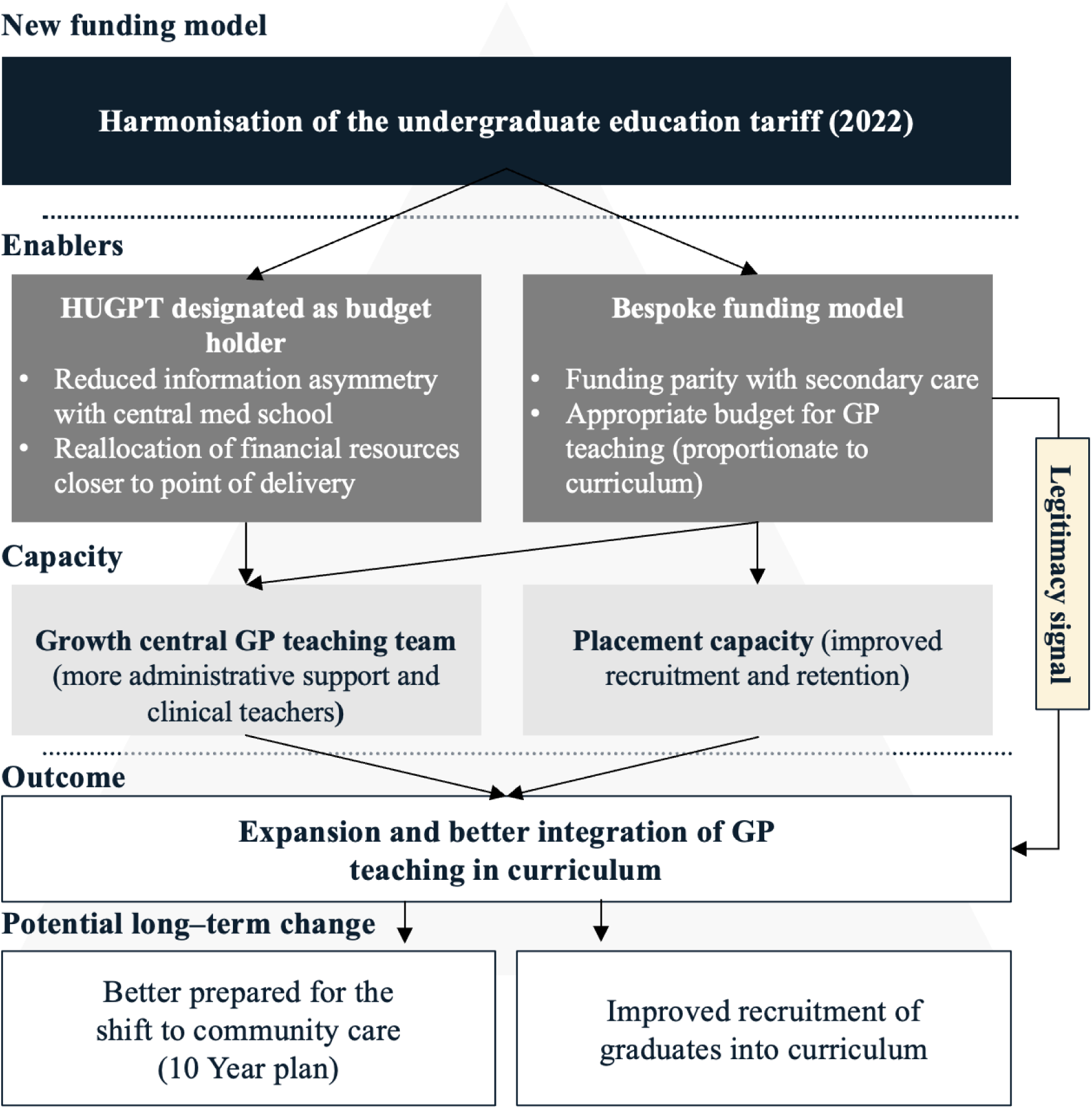
Shows the mechanisms by which the harmonised undergraduate education tariff improved the coordination and delivery of undergraduate GP teaching (figure created by authors)

Previously, there were concerns that the urgent recommendations from the *Wass* report [4] (e.g. equitable funding for GP placements and fuller curricular integration) were becoming ‘*static articles of faith*’ [23, p.164], but our findings indicate progress. The harmonised tariff represents a significant step in countering the legacy of the (Flexner-influenced) model of medical education [30], which prioritised hospital-based, subspecialty-focused training and inadvertently devalued generalism and community-based care [31], while simultaneously challenging the persistent hidden curriculum [4,18–19]. The harmonisation of the tariff functioned as a *legitimacy signal* that reframes undergraduate GP education as core rather than peripheral within medical schools [32–33].

Looking ahead, concerns were raised about future placement capacity with the increase in medical student intakes, the opening of new medical schools [25–28] and the growing numbers of postgraduate GP trainees and allied healthcare professionals. The national HUGPT group could play an important coordinating role by aligning payment rates and placement expectations across medical schools.

### Strengths and limitations

Although the sample (n=12) was small, participants represented ∼30% of the total eligible sample and achieved theme saturation. As designated budget holders, HUGPTs are a key stakeholder group and the most directly placed to describe changes following the tariff and, therefore, an important first group to research. Further research should explore other perspectives about this topic and, in due course, quantitative data to assess impacts on placement capacity, faculty development, curriculum contribution, and graduate career outcomes.

### Implications for policy

This analysis provides important insights into how changes in funding models impact the coordination, practice and delivery of clinical education. While the design and implementation of the harmonised tariff have been complex and taken considerable time and effort across medical schools [1,6], it clearly represents an excellent step towards better delivery of undergraduate medical education for students. Not only has the harmonised tariff delivered material shifts to support the delivery of changes in undergraduate medical education, but it has also begun to influence subtle yet important elements of the hidden curriculum. These changes may influence career intentions and better prepare graduates for the NHS’s shift [14] towards a more integrated, community-based model of care. Thus, by achieving funding parity between primary and secondary care, the tariff creates the necessary conditions for a curriculum that aligns more closely with the future needs of the health system and the patients it serves. As responsibilities transfer from NHSE to DHSC, recommendations for future practice based on our theoretical insights include:

The findings may have relevance for secondary care where centralised decision-making has reduced transparency and accountability for those delivering teaching [27–28]. Secondary care could bring budget ownership closer to delivery by naming accountable leads at placement block or directorate level. Furthermore, secondary care could strengthen its governance by reinforcing the equivalent provisions in Annex B [34] which require NHS placement providers to demonstrate that tariff funding is used for placement delivery.

Finally, while greater local oversight may support transparency, it is important not to lose the involvement of medical schools in primary care tariff governance. In contrast to secondary care, where funding now flows directly from NHS England to hospitals with little university input, the primary care model benefits from the combined educational and practical expertise of universities and teaching practices.

## Supporting information

Supplementary File 1: SRQR reporting checklist

## Data Availability

All data produced in the present study are available upon reasonable request to the authors

## Acknowledgments

We thank all the HUGPTs who volunteered and took part in the study, as well as the SAPC HUGPT group. We are grateful to Professor Joe Rosenthal for his guidance and helpful suggestions.

## Ethical approval

This study was conducted in accordance with the Declaration of Helsinki and institutional research governance standards. The University of Oxford Medical Sciences Interdivisional Research Ethics Committee (MS IDREC) reviewed the project and determined that it constituted a service evaluation, and therefore did not require formal ethical approval. No patient or identifiable clinical data were collected. Data was handled and stored securely on encrypted University servers in line with University policies.

## Human Ethics and Consent to Participate declarations

All participants received an information sheet, provided verbal informed consent (including consent to record), and took part voluntarily.

