## Supplementary File 1: SRQR reporting checklist for "The Impact of the Harmonised Education Tariff on Faculty Development and Teaching Capacity in Undergraduate GP Education: A Qualitative Interview Study"

### Standards for Reporting Qualitative Research (SRQR)

O'Brien B.C., Harris, I.B., Beckman, T.J., Reed, D.A., & Cook, D.A. (2014). Standards for reporting qualitative research: a synthesis of recommendations. *Academic Medicine, 89(9)*, 1245-1251.

| No. Topic | Item |
|---|---|
| **Title and abstract** | |
| S1 Title | The title clearly identifies the study topic and design "a *qualitative interview study"* and specifies the focus: *impact of the harmonised education tariff on undergraduate GP teaching*. |
| S2 Abstract | Structured abstract following BJGP format, summarising objective, design, setting, participants, methods, results (six themes), and conclusion on the tariff's organisational and educational impact. |
| **Introduction** | |
| S3 Problem formulation | Situates the problem in the long-standing funding inequity between primary and secondary care placements, referencing SIFT and its limitations. Synthesises policy history, the Wass Report, and existing evidence gaps. Establishes significance: underfunding threatens GP exposure and recruitment. |
| S4 Purpose or research question | To examine how the harmonised undergraduate tariff has impacted the coordination, delivery, and sustainability of undergraduate GP teaching across England. |
| **Methods** | |
| S5 Qualitative approach and research paradigm | Adopts a *reflexive thematic analysis* (Braun & Clarke) within a *constructivist–interpretivist* paradigm. Theoretical framing integrates *Principal–Agent Theory*, *Resource Dependence Theory*, and *Institutional Theory* to interpret organisational change. |
| S6 Researcher characteristics and reflexivity | Conducted by a DPhil researcher in primary care education and a senior GP academic. The research team's insider knowledge (education policy, GP teaching) informed data interpretation. Reflexive awareness maintained through team discussions and |

| | |
|---|---|
| | iterative coding; influence acknowledged as shaping lens rather than biasing findings. |
| S7 Context | Study conducted within UK medical education; setting: twelve medical schools across England and two from devolved nations. Context shaped by recent policy shift (2022 tariff) and transition from NHSE to DHSC. |
| S8 Sampling strategy | Purposive sampling of *Heads of Undergraduate GP Teaching* (HUGPTs), representing institutional and geographical diversity. Sampling ceased at *thematic saturation* (n=12), judged sufficient for analytic depth and variation. |
| S9 Ethical issues pertaining to human subjects | Classified by University of Oxford MSD IDREC as *service evaluation*; formal ethics review not required. Informed consent obtained verbally and recorded. Data anonymised, stored on encrypted servers, adhering to GDPR. |
| S10 Data collection methods | Semi-structured interviews (30–60 minutes) via Microsoft Teams, June–August 2025. Iterative approach: topic guide refined as themes emerged. Data collection ended after saturation. |
| S11 Data collection instruments and technologies | Interview topic guide (Box 1) informed by literature and early insights. Microsoft Teams used for recording. Minor adjustments made to prompts during data collection to explore emergent issues. |
| S12 Units of study | Twelve HUGPTs across 12 UK medical schools (mix of Russell Group, established, and new schools). Participants varied by role tenure, institutional type, and curriculum model. |
| S13 Data processing | Interviews transcribed verbatim, anonymised, checked for accuracy. Managed in NVivo 15. Secure storage on University servers. |
| S14 Data analysis | Inductive reflexive thematic analysis following Braun & Clarke's six-phase process. Coding and theme generation iterative, guided by the research question and evolving insights. Theoretical interpretation mapped findings to organisational theories |
| S15 Techniques to enhance trustworthiness | Multiple readings, iterative coding, analytic memos, and team discussions ensured reflexivity and coherence. OSF pre-registration enhanced transparency; full topic guide and coding tree shared as supplementary files. |
| **Results/Findings** | |
| S16 Synthesis and interpretation | |

| | |
|---|---|
| | Six overarching themes identified: budget control and organisational change; curriculum expansion; placement capacity; teaching quality; student experience; accountability frameworks. Interpretation highlights mechanisms linking tariff reform to structural change. |
| S17 Links to empirical data | Findings substantiated with anonymised quotes throughout the Results, illustrating variation and consensus across participants. |
| **Discussion** | |
| S18 Integration with prior work, implications, transferability, and contribution(s) to the field | Findings extend Rosenthal et al. (2022) by offering first empirical evaluation of the tariff's impact. Integrates with policy literature on equity and GP recruitment. Theorised contribution: illustrates how funding parity shifts organisational power, legitimacy, and curriculum capacity. Transferability discussed to similar systems facing funding asymmetry. |
| S19 Limitations | Acknowledges small but diverse sample; reliance on HUGPT perspectives; absence of triangulation with tutors/students. Calls for future quantitative and mixed-method studies. |
| **Other** | |
| S20 Conflicts of interest | None declared. |
| S21 Funding | Supported by MRC–Optum Industrial CASE Studentship (MR/W006731/1). Funders had no role in data collection, analysis, or interpretation. |
